# Forecasting Dengue across Brazil with LSTM Neural Networks and SHAP-Driven Lagged Climate and Spatial Effects

**DOI:** 10.1101/2024.12.11.24318832

**Authors:** Xiang Chen, Paula Moraga

**Author notes:** Contributing authors.

## Abstract

**Background:** Dengue fever is a mosquito-borne viral disease that poses significant health risks and socioeconomic challenges in Brazil, necessitating accurate forecasting across its 27 federal states. With the country’s diverse climate and geographical spread, effective dengue prediction requires models that can account for both climate variations and spatial dynamics. This study addresses these needs by using Long Short-Term Memory (LSTM) neural networks enhanced with SHapley Additive exPlanations (SHAP) integrating optimal lagged climate variables and spatial influence from neighboring states.

**Method:** An LSTM-based model was developed to forecast dengue cases across Brazil’s 27 federal states, incorporating a comprehensive set of climate and spatial variables. SHAP was used to identify and select the most important lagged climate predictors. Additionally, lagged dengue cases from neighboring states were included to capture spatial dependencies. Model performance was evaluated using MAE, MAPE, and CRPS, with comparisons to baseline models.

**Results:** The LSTM-Climate-Spatial model consistently demonstrated superior performance, effectively integrating temporal, climatic, and spatial information to capture the complex dynamics of dengue transmission. SHAP-enhanced variable selection improved accuracy by focusing on key drivers such as temperature, precipitation and humidity. The inclusion of spatial effects further strengthened forecasts in highly connected states showcasing the model’s adaptability and robustness.

**Conclusion:** This study presents a scalable and robust framework for dengue forecasting across Brazil, effectively integrating temporal, climatic, and spatial information into an LSTM-based model. By integrating diverse data sources, the framework captures key transmission drivers, demonstrating the potential of LSTM neural networks for robust predictions. These findings provide valuable insights to enhance public health strategies and outbreak preparedness in Brazil.

## 1 Introduction

Dengue fever, a mosquito-borne viral disease caused by the dengue virus (DENV), remains one of the most pressing public health issues globally, posing a major health threat to half of the world’s population [1, 2]. The dengue virus is primarily transmitted to humans by the *Aedes aegypti* mosquito, which has adapted well to urban environments, making the virus easily transmissible in densely populated areas [3, 4]. The disease, endemic to more than 100 countries, has seen a dramatic surge in recent decades, with incidence rates increasing thirty-fold over the last 50 years [5]. The World Health Organization (WHO) estimates that dengue infections now number around 390 million annually, with approximately 3.9 billion people worldwide at risk of infection [1].

In Brazil, dengue’s history can be traced back to the late 19th century, with the first major recorded outbreak occurring in Boa Vista, Roraima, in 1981 [6]. Despite eradication efforts targeting *Aedes aegypti* in the 1950s, the mosquito was reintroduced in the 1970s, leading to regular epidemics throughout the country [7]. The year 2024 marked a historic dengue outbreak in Brazil, with approximately 6.6 million probable cases and 5,867 deaths reported by epidemiological week 48 (first week of December), 2024 [8]. This unprecedented epidemic reflects an expansion of dengue into previously unaffected regions, such as Brazil’s southern municipalities, which reported significant increases in cases for the first time [9]. These trends highlight the pressing need for robust forecasting models to guide public health interventions.

Brazil, characterized by its vast and varied climate, provides an environment conducive to the mosquito’s proliferation, enabling dengue’s persistent transmission. Consequently, Brazil consistently ranks among the nations with the highest dengue incidence worldwide [10, 11]. A confluence of factors contributes to Brazil’s vulnerability, namely, the favorable tropical climate, urbanization, demographic changes, and socio-environmental conditions that support the mosquito’s lifecycle and virus transmission [12].

There are four distinct but antigenically related serotypes of the dengue virus (DENV-1 to DENV-4), each of which has circulated extensively across Brazil [13, 14]. Infection with one serotype confers lifelong immunity to that specific type but only partial and short-lived immunity to others, making secondary infections common and often more severe, potentially resulting in dengue hemorrhagic fever (DHF) or dengue shock syndrome (DSS) [15]. In Brazil, major epidemics have been linked to the sequential circulation of these serotypes, with DENV-2 and DENV-3 frequently associated with severe cases [16]. The presence of all four serotypes and the co-circulation of these strains contribute to Brazil’s high morbidity rates and the cyclical nature of its epidemics.

The impact of climate on dengue transmission is complex. Factors such as temperature, precipitation, and humidity play crucial roles in the lifecycle of *Aedes* mosquitoes and the virus’s transmission [17]. Warmer temperatures accelerate the mosquito’s breeding cycle, decrease the time required for the virus to mature within the mosquito, and increase the feeding frequency, all of which amplify transmission rates [18]. However, the relationship between temperature and transmission is nonlinear, with an optimal range for dengue virus transmission. Studies show that transmission by *Aedes aegypti* peaks at temperatures between 26°C and 29°C, and declines sharply below 18°C and above 34°C, where mosquito survival and virus incubation rates are severely impacted [19, 20]. These nonlinear thermal thresholds highlight the need for precise, data-driven models that account for nonlinear climate responses to accurately predict the impact of climate on dengue transmission. Brazil’s climatic diversity—from the hot and humid Amazon basin to the cooler southern regions—results in varied dengue incidence rates across the country. Seasonal patterns also influence dengue outbreaks, with higher incidences observed during the rainy season (December to May) when mosquito populations thrive [18]. Despite seasonal predictability, the interplay between climate and dengue transmission is further complicated by Brazil’s geographic, demographic, and socio-economic diversity, creating a complex landscape for disease forecasting and intervention [21].

Given the high disease burden and the geographic spread of dengue across Brazil, there is an urgent need for accurate, localized prediction models to support public health planning [22]. Current prevention strategies primarily target the mosquito vector, as there remains no effective antiviral treatment for dengue. In recent years, efforts to predict dengue outbreaks have increasingly incorporated climate and spatial data to improve prediction accuracy. However, the scope of these studies often remains limited, either focusing on specific cities or states or employing models that do not adequately capture the complex spatial-temporal dynamics at play.

Forecasting dengue cases has seen the application of diverse methodologies, blending statistical models, machine learning techniques, and the integration of external factors like climate and spatial data. Traditional models such as ARIMA and SARIMA have long been utilized for their ability to capture trends and seasonal patterns in time-series data, often leveraging lagged case data to improve short-term predictions. Studies in Brazil [23, 24], Thailand [25], and other regions [26] have demonstrated their effectiveness, particularly for straightforward outbreak timing and intensity forecasts. However, these models face limitations in handling non-linear relationships or unexpected changes, which are common in dengue dynamics influenced by external environmental or social factors.

To address these challenges, machine learning methods have emerged as flexible tools capable of modeling complex, non-linear interactions. Techniques such as Random Forest, Support Vector Machines (SVM), and neural networks, particularly Long Short-Term Memory (LSTM) networks, have shown great promise in dengue prediction [27]. These models can learn intricate temporal dependencies and patterns in disease spread, often outperforming traditional statistical approaches when sufficient data are available. Hybrid architectures, combining convolutional and recurrent components, further enhance predictive power by capturing both temporal and spatial dynamics. Applications of these models across regions such as Brazil [28], India [29], Colombia [30] and Malaysia [31] have highlighted their ability to integrate diverse predictors, such as climate, socio-demographics, and historical case data, to provide accurate forecasts over different time horizons.

The inclusion of climate and environmental data has been pivotal in improving the accuracy of both statistical and machine learning approaches. Variables like temperature, rainfall, and humidity, which directly influence mosquito breeding and survival, are critical predictors of dengue transmission. Both traditional statistical and machine learning models have benefited from this integration, as these climatic factors provide critical context for understanding disease spread. For instance, statistical models incorporating climate data have been shown to outperform purely case-based models in regions like Guadeloupe [32], Mexico [33], Brazil [34], and Myanmar [35], while machine learning models integrating environmental variables have delivered improved long-term predictions in Malaysia [36], and Colombia [37].

The temporal relationship between climatic factors and dengue occurrence is critical, as it affects mosquito vector dynamics and virus transmission over time, leading researchers to apply various lags when predicting dengue. [38] observed that dengue outbreaks in Noumea, New Caledonia, lagged 1–2 months behind peak temperatures, aligning with maximum precipitation and humidity. [39] found that monthly minimum temperature with a one-month lag and cumulative precipitation with a three-month lag were effective predictors of dengue in Guangzhou, China. [32] identified minimum temperature at a lag of five weeks as a significant predictor in Guadeloupe, French West Indies. [23] showed that lag-0 maximum temperature and lag-1 rainfall correlated with dengue cases in Rio de Janeiro, Brazil. [40] reported that temperature and humidity at a two-month lag were significant predictors of dengue trends in Taiwan.

In addition to climate variables, spatial factors have been widely recognized as crucial in improving the accuracy of dengue prediction models. Studies have shown that incorporating neighboring regions’ dengue case data significantly enhances model performance [41]. For instance, ensemble ARIMA models integrating spatial effects outperformed traditional ARIMA models by accounting for dengue patterns in adjacent districts, as demonstrated in Selangor, Malaysia [42]. Similarly, research in southern Vietnam revealed significant spatial correlations in epidemic timing and magnitude within districts located 50–100 km apart, emphasizing the role of local drivers like microclimatic conditions and human mobility [43]. At finer scales, cluster analyses in Thai villages identified focal dengue transmission within 100 meters of index cases, where localized factors such as mosquito density and human movement patterns played a pivotal role [44]. Moreover, temporal correlations across neighboring areas in Taiwan have been effectively utilized to predict outbreak risks [45]. These findings collectively demonstrate that incorporating spatial dependencies into predictive models provides a more comprehensive understanding of dengue spread, enabling a more comprehensive understanding of disease dynamics and enhancing the precision of predictive frameworks.

In this study, we propose an enhanced Long Short-Term Memory (LSTM) neural network model [46, 47] to predict dengue cases at the state level across Brazil. The model integrates a diverse set of lagged climate variables, including temperature, relative humidity, precipitation, atmospheric pressure, thermal range, and rainy days, to account for environmental factors influencing dengue transmission. To optimize model performance, we utilize SHapley Additive exPlanations (SHAP) [48], which identify and select the most critical variables and their lag structures, ensuring that the model focuses on the most impactful predictors.

Incorporating a spatial component, the model also leverages lagged dengue cases from neighboring states, capturing the spatial dependencies and cross-regional influences critical to understanding disease dynamics. Additionally, seasonal patterns are included to reflect the cyclic nature of dengue outbreaks. Our approach employs a moving window strategy, using fixed 7-year periods (2016–2022) to predict dengue cases for 2023, with forecasts generated for 4 weeks (1 month) and 12 weeks (3 months) ahead. This scalable framework offers robust, adaptable state-level predictions across diverse geographical and climatic contexts in Brazil (Figure 1).

**Fig. 1.**
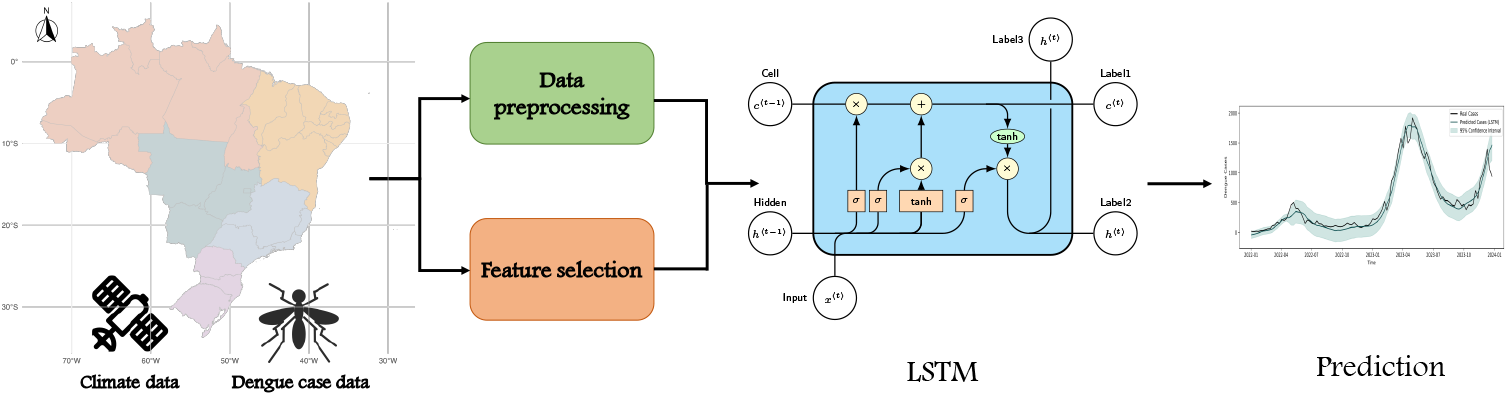
Data processing pipeline for the forecasting of dengue cases in Brazil.

To evaluate the model’s performance, we compare it against three alternative models, including simpler LSTM models and a Bayesian hierarchical model baseline. Specifically, we use a LSTM using only dengue case data; a LSTM with lagged climate variables and seasonal patterns (without neighbor spatial effects); and a Bayesian random effects model using the same covariates as our proposed model. Model performance is assessed using three metrics - Mean Absolute Error (MAE), Mean Absolute Percentage Error (MAPE), and Continuous Ranked Probability Score (CRPS) - to comprehensively evaluate accuracy, reliability, and uncertainty in the forecasts.

## 2 Methods

### 2.1 Study design

We develop an LSTM model for dengue forecasting in Brazil by combining SHAP-selected climate variables, spatial effects from neighboring states, cyclic patterns, and historical dengue case data. Forecasts are made at the state (federation unit) level, enabling the generation of insights for all 27 federal units, with the flexibility to adapt the approach to specific regional contexts. Focusing on medium-term forecasting horizons, our model generates forecasts for two horizons: 1 month (4 weeks) and 3 months (12 weeks) ahead. Very shorter-term forecasts, such as 1-week predictions, often lack sufficient lead time to enable effective public health interventions. By concentrating on 1- and 3-month forecasts, the study aims to provide actionable insights, equipping public health authorities with timely information to prepare for and mitigate potential outbreaks more effectively.

We implemented a moving window strategy with a fixed window size of 7 years to predict dengue cases at the state level across Brazil (Figure 2). This approach was chosen to balance capturing long-term trends and seasonal patterns of dengue incidence while ensuring a sufficiently large dataset for robust model training. The initial training window spanned 2016-01-03 (the first epidemiological week of 2016) to 2022-12-25 (the last epidemiological week of 2022), comprising 364 weeks (52 weeks per year *×* 7 years). The window was then moved forward by one week at a time to generate predictions for 2023, with the process repeated iteratively until the last epidemiological week of 2023. By continuously updating the training data to include the most recent observations, this strategy enables the model to adapt dynamically to evolving temporal patterns and maintain predictive accuracy.

**Fig. 2.**
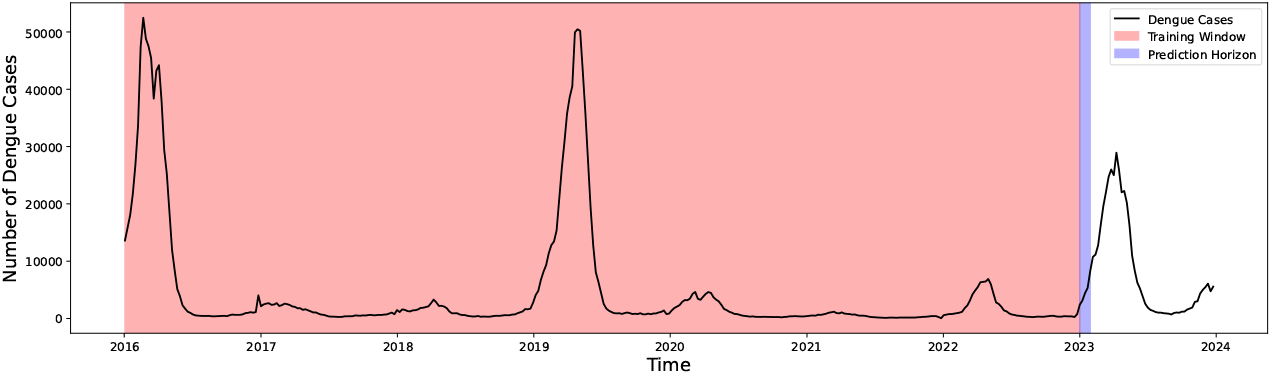
Illustration of the moving window strategy for dengue case prediction.

### 2.2 Study site

Brazil, the largest country in South America, is geographically and ecologically diverse. It is bordered by ten countries and the Atlantic Ocean, covering an area of over 8.5 million square kilometers. Brazil’s population reached an estimated 212.6 million people on July 1, 2024, according to the Brazilian Institute of Geography and Statistics (IBGE), making it the most populous country in Latin America. Over 30% of the population resides in 48 cities with more than 500,000 inhabitants, with São Paulo being the most populous municipality [49].

Brazil’s diverse geography creates significant variations in climate, which directly influence dengue transmission dynamics. Figure 3 shows a state-level map of Brazil, highlighting the regional divisions. The country is divided into five macro-regions: North, Northeast, Central-West, Southeast, and South, each presenting distinct climatic, demographic, and epidemiological profiles.

**Fig. 3.**
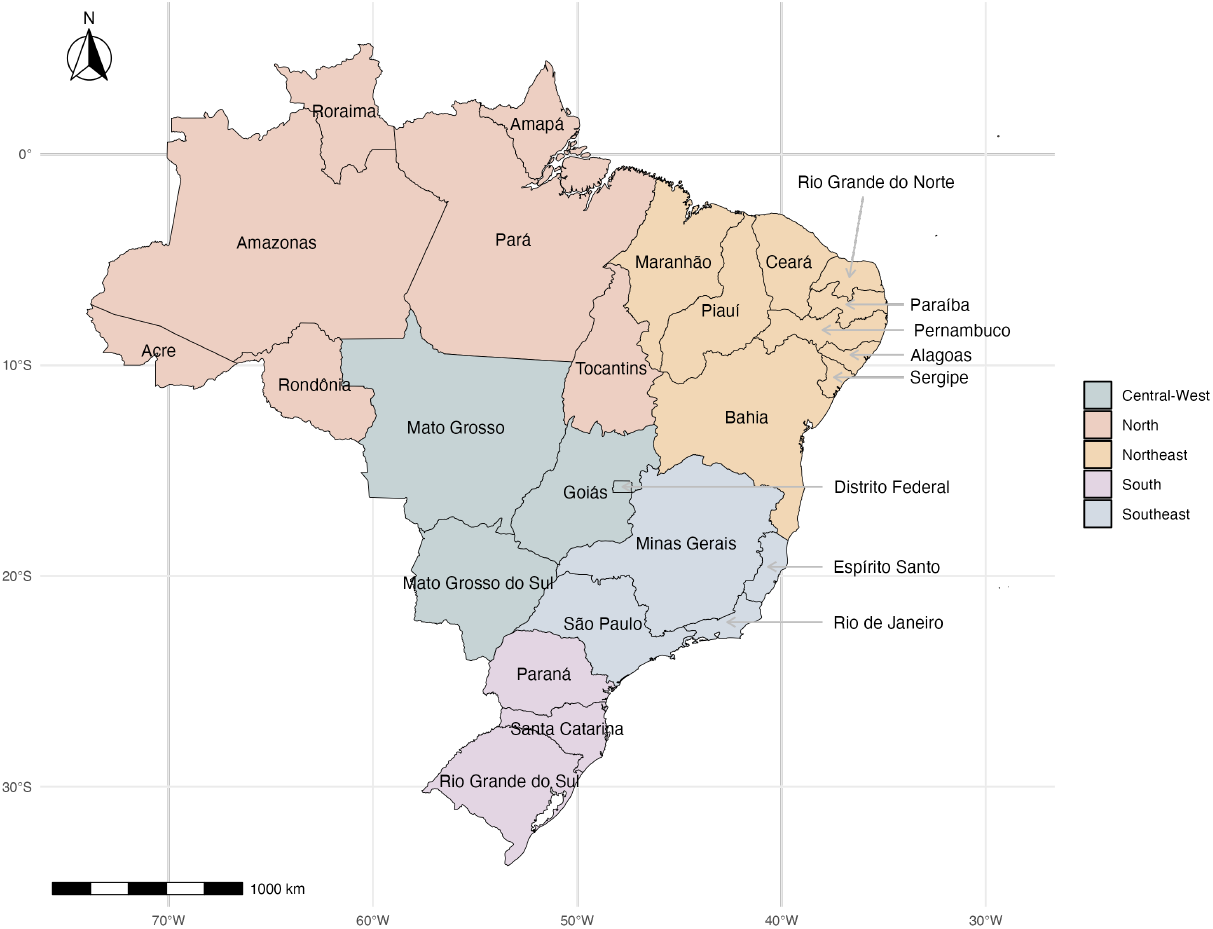
State-level map of Brazil highlighting regional divisions.

The population density varies significantly across regions, as shown in Figure 4. The Southeast region, home to São Paulo and Rio de Janeiro, is the most densely populated and economically developed area, whereas the North region, dominated by the Amazon rainforest, has sparse population density. Urbanization and human mobility patterns in densely populated regions amplify the risk of dengue transmission.

**Fig. 4.**
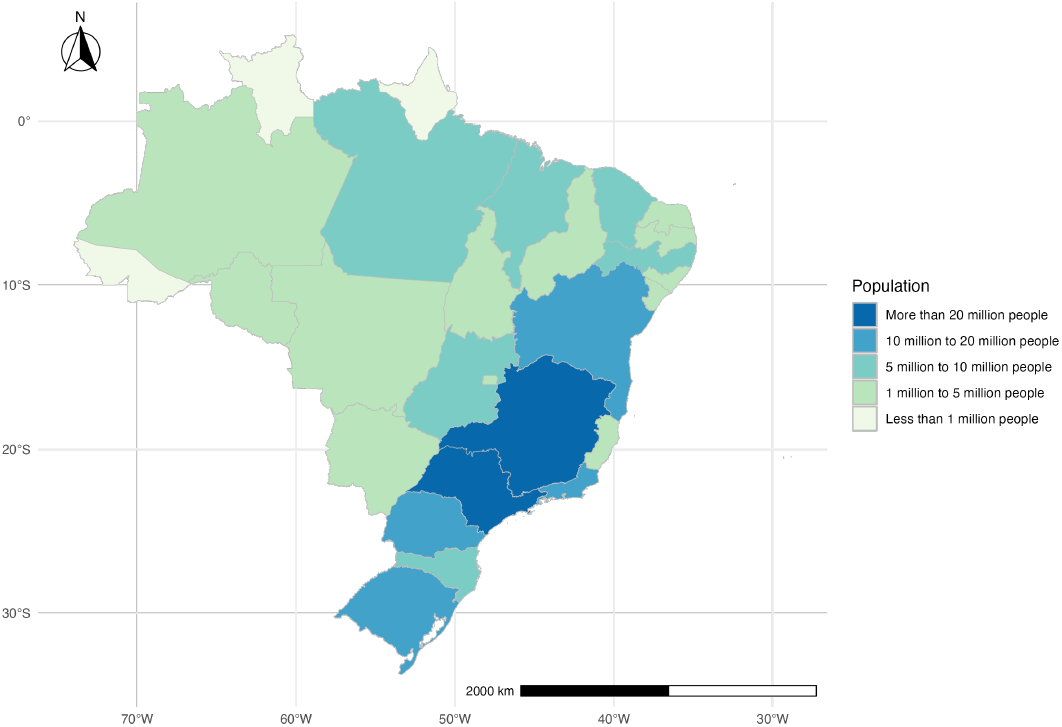
Population map in each state of Brazil from the 2022 census [51].

Brazil’s climate varies from tropical in the North to temperate in the South, with significant regional differences captured by the Köppen climate classification [50] (Figure 5). The North and Northeast regions predominantly experience tropical rain-forest and savanna climates, characterized by high temperatures and seasonal rainfall. The Central-West region, with its tropical savanna climate, has distinct wet and dry seasons, influencing the temporal dynamics of dengue outbreaks. The Southeast and South regions, with their subtropical and temperate climates, have historically reported fewer dengue cases, though recent outbreaks suggest climate change and urbanization are expanding transmission zones [9].

**Fig. 5.**
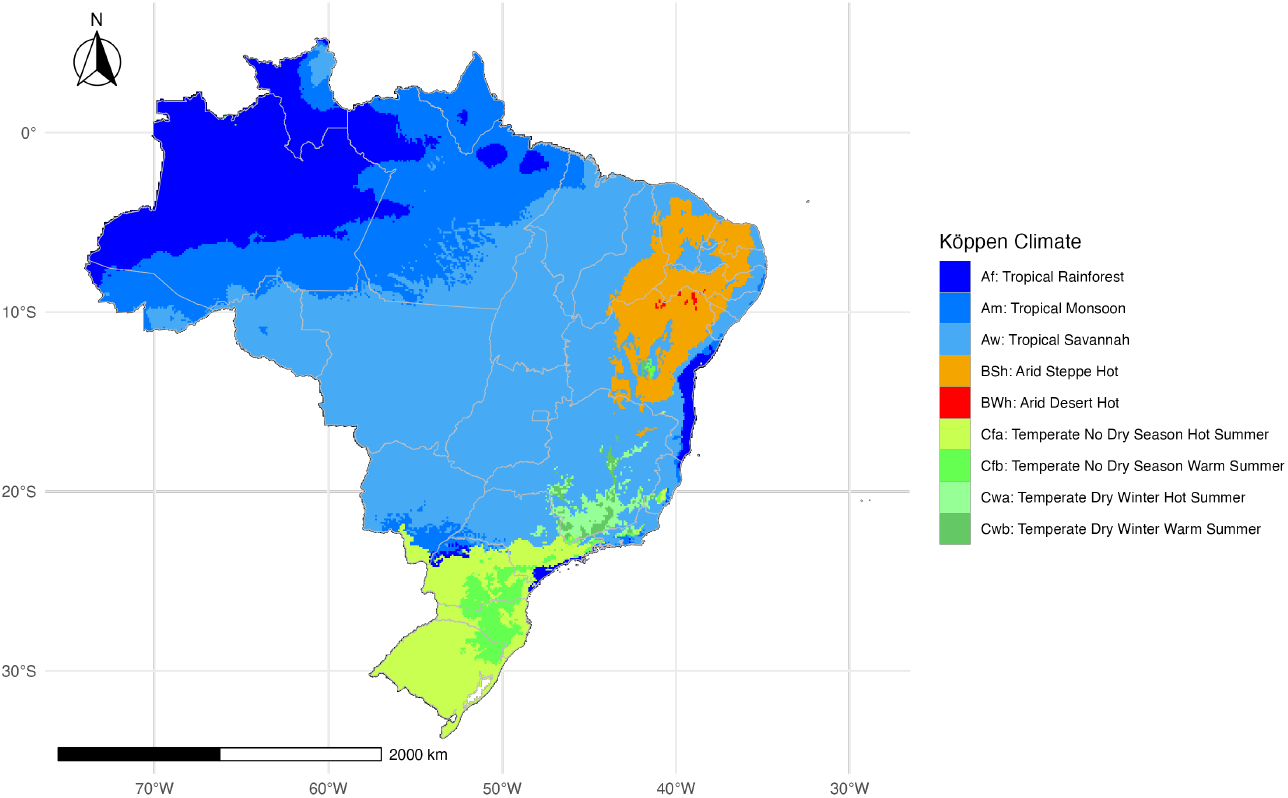
Köppen climate classification map of Brazil [50].

This diversity in climate and population density underscores the importance of developing dengue forecasting models tailored to Brazil’s unique geographical and ecological contexts.

### 2.3 Dengue data

The dengue data used in this study is sourced from InfoDengue [52], a public health surveillance system designed to monitor dengue and other arboviruses across Brazil. InfoDengue integrates official dengue case reports with meteorological data, providing a comprehensive overview of the factors driving disease transmission. The system operates in 788 cities across Brazil, offering timely and up-to-date epidemiological information to health agents, local decision-makers, and the public.

The dataset provides weekly dengue case counts for each epidemiological week, allowing for high temporal resolution in forecasting efforts. This granularity ensures that seasonal patterns and short-term trends in dengue transmission are accurately captured, forming the foundation for our medium-term prediction horizons.

Figure 6 illustrates the monthly dengue incidence rate (per 100,000 people) for each federal unit from 2016 to 2023, highlighting the geographical and temporal variability of dengue across Brazil. The states are ordered by their geographical location, emphasizing the regional heterogeneity in dengue dynamics. Northern states like Amazonas and Pará show consistent transmission patterns with peaks during the rainy season, while southern states such as Santa Catarina and Rio Grande do Sul exhibit lower and more sporadic transmission. Additionally, states like Goiás, Minas Gerais, São Paulo, Ceará, and Paraná stand out with very high incidence rates (dark red), emphasizing their vulnerability to severe outbreaks. These patterns underscore the regional heterogeneity of dengue dynamics and the necessity of incorporating climatic and spatial factors into forecasting models.

**Fig. 6.**
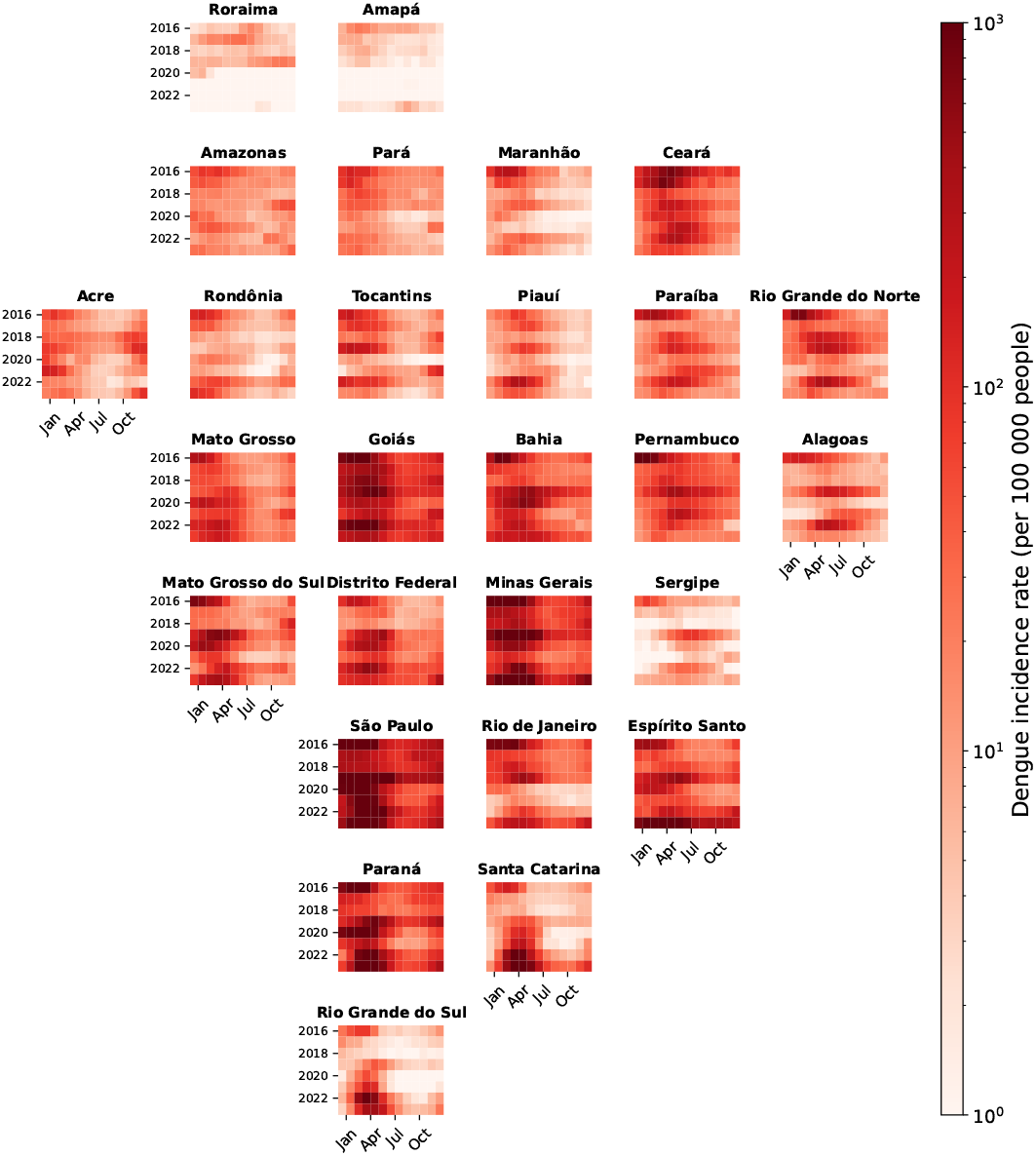
Monthly dengue incidence rate (per 100 000 people) for each federal unit between 2016 and 2023. States are ordered by their geographical location.

### 2.4 Climate data

The climate data used in this study is derived from the Copernicus ERA5 reanalysis dataset [53], which provides comprehensive global weather and climate information with high spatial and temporal resolution. This dataset includes hourly estimates of atmospheric, oceanic, and land-surface variables, making it an invaluable resource for understanding environmental factors influencing dengue transmission. To align with our epidemiological analysis, we considered ERA5 data on a weekly basis by utilizing weekly averages for each variable.

At the state level, climate data were aggregated using a population-weighted average to ensure that the derived values accurately reflect the conditions experienced by the majority of the population in each state. The population data used for weighting were obtained from the Brazilian Institute of Geography and Statistics (IBGE) [54]. This approach accounts for population density variations, providing a more representative measure of climatic influences on dengue cases across diverse regions.

Table 1 shows the climate variables considered in this study, including minimum, median, and maximum values for temperature, precipitation, and atmospheric pressure, as well as thermal range, relative humidity, total precipitation, and the number of rainy days per week. These variables capture key environmental conditions linked to mosquito survival, breeding, and dengue transmission.

**Table 1.**
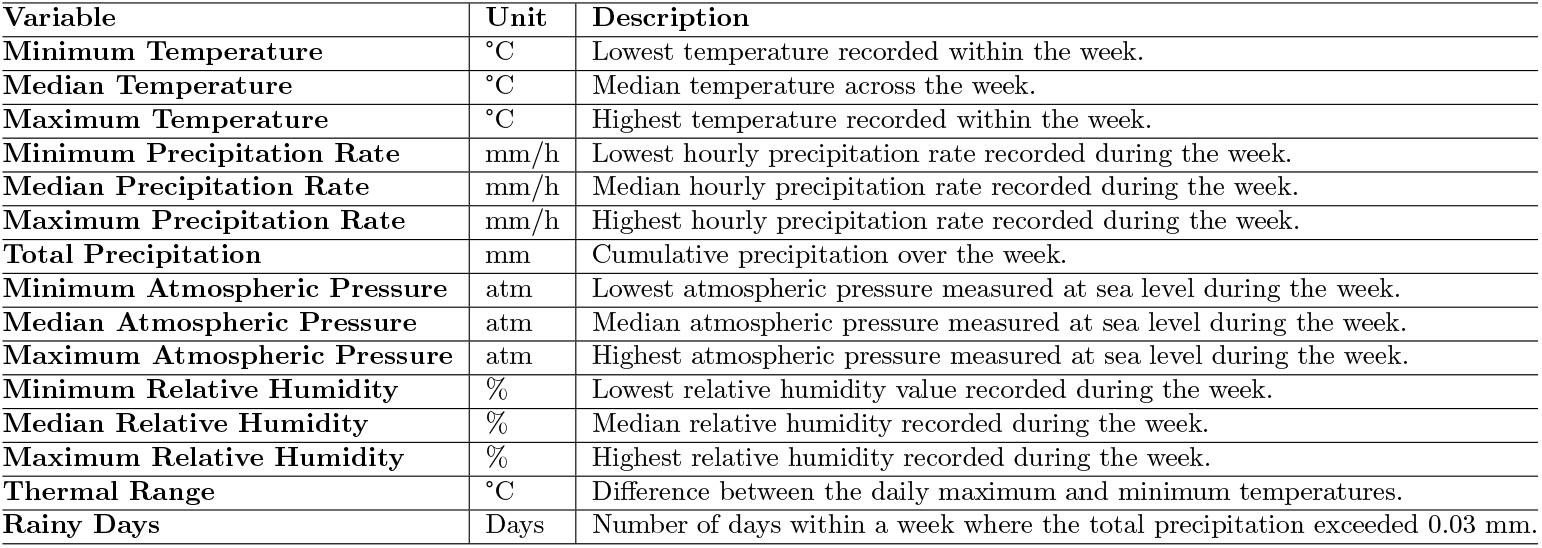
Climate Variables Derived from Copernicus ERA5 Reanalysis Data, Summarized by Week.

Climate variables are considered with a lag of 1–3 months to predict dengue cases one month (4 weeks) ahead, and a lag of 3 months for predicting three months (12 weeks) ahead. This choice aligns with findings from previous studies, and is also driven by data availability, as climate information is typically available only up to the current time, creating a natural lag when predicting future dengue cases. The selected lags effectively capture both immediate and cumulative climatic influences on dengue transmission by accounting for the temporal relationship between environmental conditions and mosquito vector dynamics.

### 2.5 Feature selection with SHAP

To enhance the interpretability and efficiency of the model, we employ SHapley Additive exPlanations (SHAP) [48] for feature selection. SHAP is a game-theoretic approach that quantifies the contribution of each feature to the model’s predictions, based on the concept of Shapley values from cooperative game theory. This method provides a principled and consistent way to attribute the prediction of the model to individual input features.

SHAP explains the output *f* (*x*) of a model for a specific input *x* = {x_1_, *x*_2_, …, *x*_*n*_} by computing the contribution of each feature *x*_*i*_. The contribution, or Shapley value, is calculated as the average marginal contribution of *x*_*i*_ across all possible feature subsets *S* ⊆ *{x*_1_, *x*_2_, …, *x*_*n*_}:

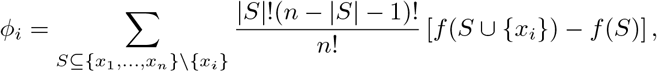

where *ϕ*_*i*_ represents Shapley value for feature *i*; *S* is a subset of features excluding *x*_*i*_; *f* (*S*) is the model prediction using only features in subset *S*; and *n* is the total number of features. This approach ensures that the contributions of all features sum to the total model output, offering a fair and consistent attribution of importance.

In this study, SHAP is used to rank the importance of climate variables derived from the Copernicus ERA5 reanalysis dataset [53]. Climate variables such as temperature, precipitation, atmospheric pressure, relative humidity, thermal range, and rainy days often show high correlations within their respective groups (e.g., minimum, median, and maximum values). SHAP identifies the most influential variables among these correlated groups, potentially selecting more than one if their contributions to the model are equally significant. This reduces redundancy while preserving predictive power and computational efficiency. For example, SHAP may retain both median and maximum temperature if they provide complementary predictive insights.

To implement SHAP-based feature selection, an initial LSTM model is trained with all candidate climate variables. This process is carried out separately for each state to account for regional differences in dengue transmission dynamics. Shapley values are then computed for each feature, and variables with the highest average absolute Shapley values are retained. By focusing on the most impactful variables, SHAP ensures that the LSTM model captures the critical relationships between climate factors, spatial effects, and dengue transmission. This systematic approach reduces the feature space while enhancing interpretability by clearly identifying the climate variables driving dengue transmission.

### 2.6 Cyclic seasonal component

Dengue transmission exhibits strong seasonal patterns, primarily influenced by climatic conditions such as temperature, precipitation, and humidity, which affect mosquito breeding and virus transmission cycles. While climatic variables are included as predictors in the model, the model also incorporates a seasonal component that captures residual cyclic trends that are not fully explained by climate data alone. These trends may arise from non-climatic factors, such as human behavior, vector control efforts, and reporting practices, which tend to repeat annually.

The seasonal component is encoded using trigonometric transformations of the epidemiological week, which effectively model the cyclical nature of dengue incidence over the course of a year. Specifically, we use Seasonality_1_ = sin (2*π ×* week*/*52) and Seasonality_2_ = cos (2*π ×* week*/*52), where week represents the epidemiological week within a year (i.e., 1 to 52). By incorporating both climate variables and the seasonal component, the model ensures that the cyclical structure of dengue incidence is captured comprehensively, aligning peaks with favorable conditions while accounting for patterns beyond the direct influence of climate.

### 2.7 Neighbouring effect

Dengue transmission is influenced not only by local factors but also by spatial dependencies, as neighboring regions often share similar environmental conditions and human mobility patterns that facilitate disease spread. Therefore, in addition to climate variables and temporal patterns, we incorporate the effect of neighboring states’ dengue cases to account for spatial dependencies to improve prediction accuracy. Specifically, the model includes the lagged dengue case counts from each state’s neighbors as predictors.

Table 2 lists the neighboring states for each federal unit, where neighbors are defined as states that share a common geographical border. For instance, as illustrated in Figure 7, the state of Minas Gerais (MG) is influenced by seven neighboring states, namely, Bahia (BA), Espírito Santo (ES), Rio de Janeiro (RJ), São Paulo (SP), Mato Grosso do Sul (MS), Goiás (GO), and Distrito Federal (DF). This information ensures that the spatial context of dengue transmission is effectively captured, enabling the model to reflect the interconnected dynamics across regions.

**Table 2.**
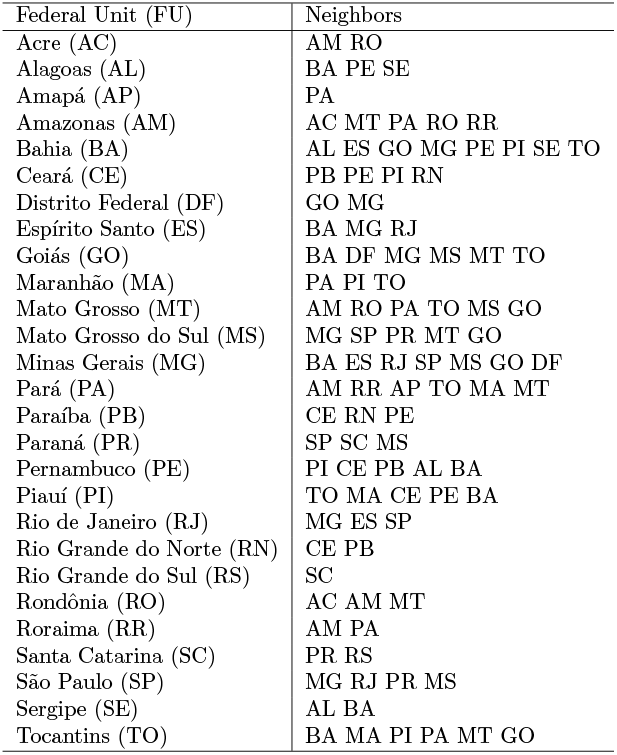
Neighbors of each of the Brazilian states.

**Fig. 7.**
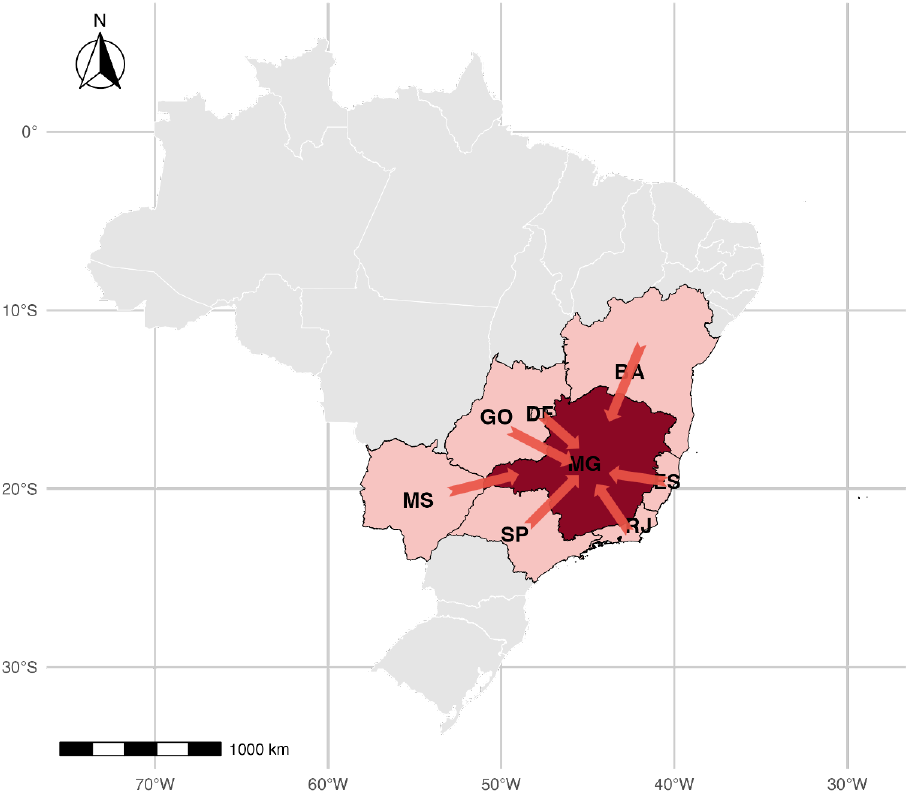
Minas Gerais (MG) and its neighboring states. Arrows indicate the spatial influence of neighbors.

### 2.8 LSTM model architecture

The forecasting framework developed in this study integrates temporal, climatic, and spatial information into a Long Short-Term Memory (LSTM) neural network. The entire process, from data preprocessing to prediction, is illustrated in the pipeline shown in Figure 1. This pipeline begins with the collection of dengue case and climate data, followed by integration of the data at the state level. Once integrated, the data undergo feature selection using SHAP, which identifies the most relevant climate variables for prediction. The refined dataset, including lagged climate variables, historical dengue cases, and neighboring states’ dengue data, is then fed into the LSTM model, which learns complex temporal patterns and spatial dependencies to generate dengue case forecasts.

The pipeline is designed to handle the intricate relationships between dengue transmission and its influencing factors, ensuring that the model captures both the immediate and broader drivers of outbreaks. This architecture combines state-level data integration, lagged variable preparation, sequential processing, and feature selection, optimizing model interpretability and accuracy.

The LSTM was chosen for its effectiveness in capturing temporal dependencies in sequential data, such as dengue case time series. The architecture consists of multiple layers designed to process both climate variables and spatial information from neighboring states described as follows.

### Input layer

The input layer receives a multivariate time series, represented as:

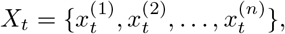

where *X*_*t*_ is the input at time *t*, and 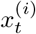 represents the *i*-th variable at time *t*. This set includes lagged dengue cases, SHAP-selected lagged climate variables, lagged dengue cases from neighboring states and cyclic seasonal patterns. Each time series input is standardized to ensure consistent scaling and convergence during training. The input dimensions are defined as (*L, n*), where *L* represents the number of previous time steps used as input for the model, and *n* is the number of features. To ensure consistent scaling and convergence during training, all inputs are standardized by transforming each input feature to have a mean of 0 and a standard deviation of 1. This step ensures that all variables are on a similar scale, preventing features with larger numerical ranges from dominating the learning process.

### LSTM layers

The core of the model comprises stacked LSTM layers, which allow the network to learn complex temporal dependencies in the data. These layers process sequential data by selectively updating their internal states through forget, input, and output gates:

1. Forget Gate:

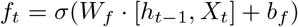

2. Input Gate:

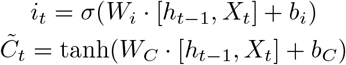

3. Cell State Update

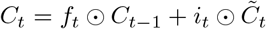

4. Output Gate

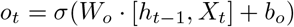

5. Hidden State Update

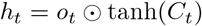

Here, *W* and *b* represent the weight matrices and bias vectors for each gate, respectively, *σ* denotes the sigmoid activation function, and ⊙ represents element-wise multiplication. The term *h*_*t*−1_ refers to the hidden state from the previous time step *t* − 1, which encodes information from earlier inputs in the sequence. These mechanisms allow the LSTM layers to retain relevant information over long time horizons while filtering out less critical details, making them highly effective for time-series data such as dengue cases. The LSTM layers are optimized to capture non-linear relationships between dengue cases, climate covariates, and spatial effects from neighboring states.

### Dense layers

Following the LSTM layers, dense (fully connected) layers are used to refine the extracted features. The output from the last LSTM cell, *h*_*T*_, is passed through the dense layers:

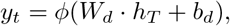

where *y*_*t*_ represents the predicted dengue cases, *W*_*d*_ and *b*_*d*_ are the weights and biases of the dense layer, and *ϕ* is the activation function (e.g., ReLU). These layers enable the model to map the learned features to the target dengue case counts.

### Output layer

The output layer is a single node that predicts the number of dengue cases for the target time step. It uses a linear activation function suitable for continuous predictions:

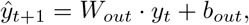

where 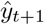 is the predicted dengue case count for the next time step.

### Regularization and optimization

To prevent overfitting, dropout is applied to the LSTM and dense layers during training:

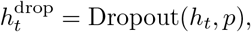

where *p* is the dropout rate. Early stopping is also used to halt training when validation performance ceases to improve. The model is trained using the Adam optimizer with the mean squared error (MSE) loss function:

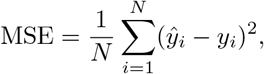

where *N* is the number of samples, *ŷ*_*i*_ is the predicted value, and *y*_*i*_ is the actual dengue case count.

### 2.9 Baseline: Bayesian baseline random effects model

As a baseline for comparison, we employ a Bayesian random effects model to predict dengue cases. This model captures temporal and spatial variability in dengue incidence by incorporating weekly counts, climate variables, cyclic seasonal patterns, and spatial effects from neighboring regions. The model is implemented using the integrated nested Laplace approximations (INLA) framework [55]. The model assumes the number of dengue cases at time *t* and region *r*, denoted as *Y*_*t*,*r*_, follows a negative binomial distribution to handle overdispersion:

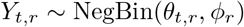

Here, *θ*_*t*,*r*_ is the mean number of cases, and *ϕ*_*r*_ is the dispersion parameter specific to each region *r*. The linear predictor log(*θ*_*t*,*r*_) includes terms for baseline effects, seasonal and weekly variations, climate influences, and spatial dependencies as follows:

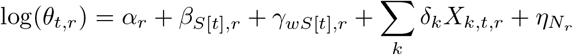

Here, the intercept *α*_*r*_ represents the baseline incidence level in region *r*, accounting for regional differences. Seasonal variations, *β*_*S*[*t*],*r*_, are included to capture region-specific seasonal patterns across epidemiological seasons *S*[*t*]. The terms *γ*_*wS*[*t*],*r*_ capture weekly cyclic variations within each season using Bayesian splines. Climate variables *X*_*k*,*t*,*r*_ such as temperature, precipitation, and humidity, consistent with those used in the LSTM model, are included to account for environmental influences on dengue transmission. Finally, the spatial effects 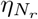, capture correlations among neighboring regions using an Intrinsic Conditional Autoregressive (ICAR) structure [56].

For each state, posterior predictive distributions are generated to forecast weekly dengue case counts. This baseline serves as a robust benchmark, incorporating the same covariates and spatial effects to evaluate the added value of our proposed model.

### 2.10 Adaptive conformal prediction

To quantify the uncertainty of dengue case forecasts, we implement an adaptive conformal prediction framework [57–60]. Conformal prediction is a robust statistical method that provides prediction intervals with a predefined confidence level, ensuring that the true value falls within the interval with a specified probability. This approach is particularly useful in epidemiological forecasting, where understanding prediction uncertainty is critical for informed public health decision-making.

For each time *t* and forecast horizon *h*, the nonconformity score is defined as the residual at time *t* for *h* steps ahead calculated as the difference between the actual and predicted values from the time series model:

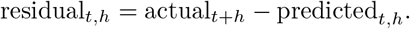

For a window size *W* and time *t*, residuals are considered from *t* − *W* to *t*. Thus, the set of nonconformity scores within the window at time *t* for a forecast horizon *h* is

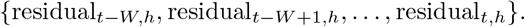

The quantiles of these nonconformity scores are then computed to determine the prediction intervals. Specifically, the lower and upper bounds of the prediction interval at each time step *t* for forecast horizon *h* are given by

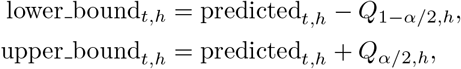

where *Q*_1−*α/*2,*h*_ and *Q*_*α/*2,*h*_ are the quantiles of the residuals within the window for the *h*-step ahead forecast, and *α* is the significance level (e.g., 0.05 for a 95% prediction interval). For the *k*-th quantile at forecast horizon *h*, the corresponding value is given by

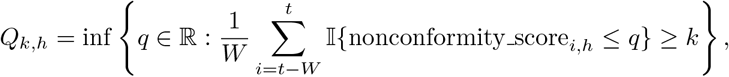

where 𝕀 is the indicator function.

The prediction interval is then adjusted dynamically at each time step based on these scores. The data is divided into training and calibration sets. The LSTM model is trained on the training set, while the calibration set is used to compute nonconformity scores. As predictions are made, the calibration set is dynamically updated to incorporate new data, allowing the prediction intervals to adapt to temporal changes in dengue transmission.

### 2.11 Forecasting accuracy

To evaluate the predictive performance of the proposed model, we use three key metrics: Mean Absolute Error (MAE), Mean Absolute Percentage Error (MAPE), and Continuous Ranked Probability Score (CRPS). These metrics collectively provide a comprehensive assessment of both the accuracy and the reliability of the forecasts.

Let *y*_*i*_ and 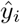 represent, respectively, the actual and forecast number of dengue cases, and let *n* be the number of observations. The Mean Absolute Error (MAE) measures the average magnitude of the errors in a set of predictions, without considering their direction. It is calculated as the average of the absolute differences between the forecasted values and the actual values:

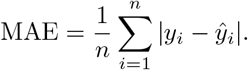

The Mean Absolute Percentage Error (MAPE) expresses the accuracy as a percentage, which provides a relative measure of the errors. It is calculated as the average of the absolute percentage differences between the predicted and actual values:

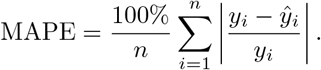

The Continuous Ranked Probability Score (CRPS) is a commonly used metric to evaluate the accuracy of probabilistic forecasts, especially for cases where we want both a precise prediction and an estimate of its uncertainty. CRPS assesses how well the forecast distribution captures the observed values. Lower CRPS values indicate better alignment between the forecasted distribution and actual outcomes, while higher values reflect poorer alignment. CRPS is calculated as follows:

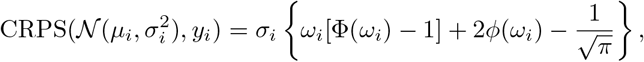

where Φ(*ω*_*i*_) and *ϕ*(*ω*_*i*_) are the cumulative distribution function (CDF) and the probability density function (PDF) of the standard normal distribution, respectively, evaluated at the normalized prediction error 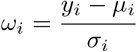. Additionally, *y*_*i*_ represents the cases observed in week *i, µ*_*i*_ is the predicted mean dengue case value for week *i*, and *σ*_*i*_ is the standard deviation of the forecast on week *i*, approximated as the length of the adaptative confidence intervals derived from the residuals between observed and predicted values.

## 3 Results

### 3.1 Feature importance

Correlation analysis among climate variables revealed similar patterns across states, with high correlations observed within variable groups, such as minimum, median, and maximum temperature, and among precipitation metrics. Figure 8 shows a heatmap for the state of Minas Gerais as an example. Given these findings, we employed SHapley Additive exPlanations (SHAP) to identify and select the most relevant variables, ensuring that only the most impactful predictors were included in the model, reducing redundancy and enhancing computational efficiency. The feature importance analysis is conducted independently for all 27 federal units of Brazil, as each state has unique climatic and epidemiological characteristics, resulting in distinct sets of influential variables.

**Fig. 8.**
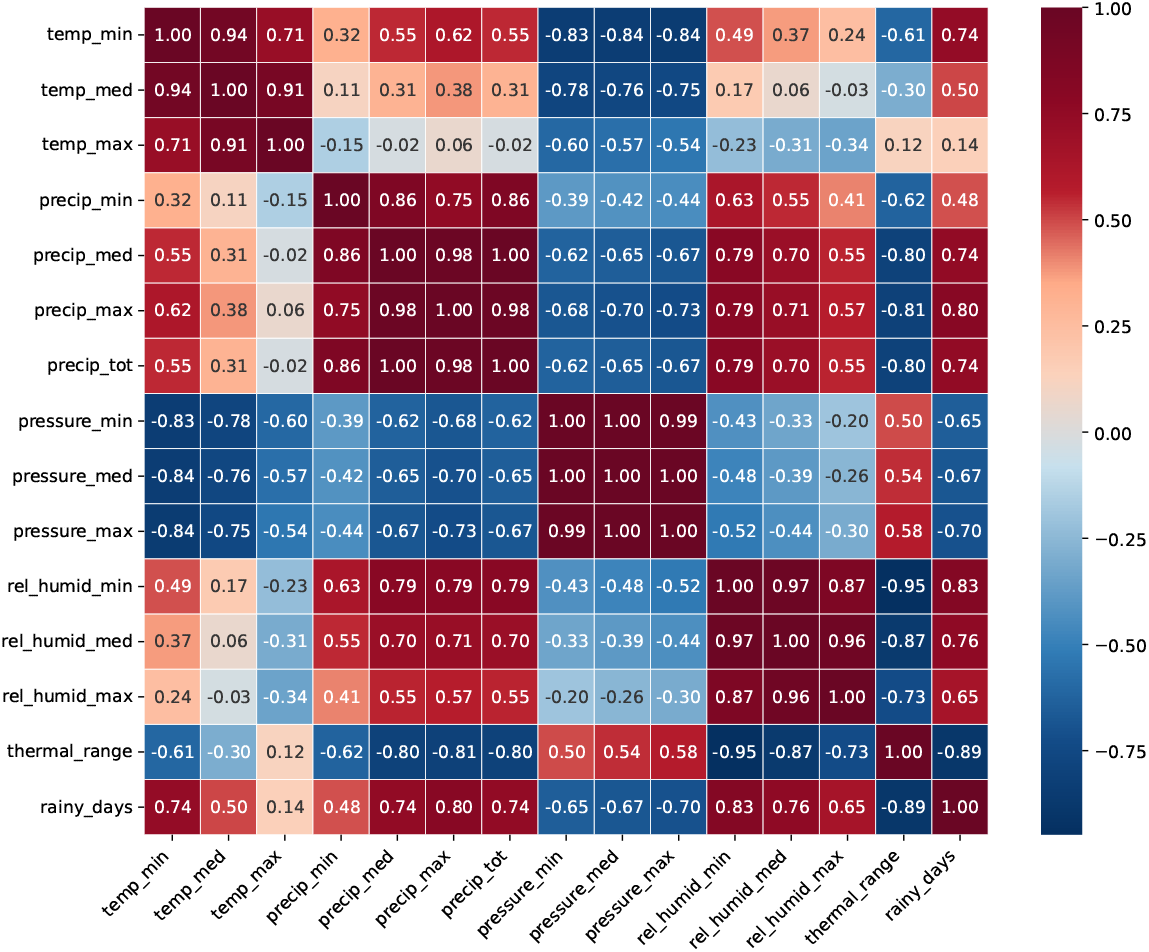
Correlation heatmap of climate variables for Minas Gerais, Brazil.

Table 3 shows the top five most important climate variables identified by SHAP for predicting dengue cases across Brazilian states. Figure 9 illustrates the SHAP summary plot for Minas Gerais as an example. Each point in the plot corresponds to a weekly prediction, colored by the magnitude of the feature values (red for high values and blue for low values). The variables are ranked by their mean absolute SHAP values, indicating their average contribution to the model’s predictions for this state. For Minas Gerais, the most influential variables include maximum relative humidity, minimum precipitation rate, and minimum temperature, suggesting that humidity, precipitation and temperature are key drivers in the dynamics of dengue transmission in this state. Other states exhibit different rankings and influential variables due to their unique climatic and epidemiological contexts. This tailored feature selection process ensures that the model captures region-specific drivers of dengue transmission, improving prediction accuracy across diverse geographical and environmental conditions in Brazil.

**Table 3.** Top five climate variables identified by SHAP for dengue prediction across Brazilian states.

| Federal Unit (FU) | Code | Top five climate variables |
| --- | --- | --- |
| Acre (AC) | 12 | pressure_max, thermal_range, pressure_min, pressure_med, rel_humid_med |
| Alagoas (AL) | 27 | rel_humid_max, rel_humid_min, precip_min, temp_med, rel_humid_med |
| Amapá (AP) | 16 | rel_humid_max, temp_min, thermal_range, rel_humid_med, precip_min |
| Amazonas (AM) | 13 | precip_med, pressure_min, rel_humid_min, temp_med, precip_min |
| Bahia (BA) | 29 | rel_humid_max, temp_max, temp_med, pressure_min, rel_humid_med |
| Ceará (CE) | 23 | rel_humid_max, temp_med, rel_humid_med, rel_humid_min, temp_min |
| Distrito Federal (DF) | 53 | rel_humid_med, precip_max, rel_humid_max, thermal_range, pressure_max |
| Espírito Santo (ES) | 32 | precip_max, rel_humid_min, precip_med, rel_humid_max, pressure_med |
| Goiás (GO) | 52 | rel_humid_max, temp_med, precip_max, temp_max, pressure_min |
| Maranhão(MA) | 21 | rel_humid_max, temp_med, precip_max, temp_max, thermal_range |
| Minas Gerais (MG) | 51 | rel_humid_max, precip_min, precip_med, temp_min, pressure_med |
| Mato Grosso do Sul (MS) | 50 | rel_humid_med, rel_humid_max, thermal_range, pressure_min, precip_max |
| Mato Grosso (MT) | 31 | rel_humid_max, precip_min, precip_med, rel_humid_med, temp_min |
| Pará (PA) | 15 | precip_min, rel_humid_min, rel_humid_max, temp_med, temp_min |
| Paraíba (PB) | 25 | thermal_range, rel_humid_min, precip_max, rel_humid_med, temp_max |
| Pernambuco (PE) | 41 | temp_med, pressure_min, pressure_max, rel_humid_max, temp_max |
| Piauí (PI) | 26 | precip_max, precip_min, rel_humid_min, rel_humid_max, thermal_range |
| Paraná (PR) | 22 | temp_med, rel_humid_min, rainy_days, pressure_min, pressure_max |
| Rio de Janeiro (RJ) | 33 | thermal_range, temp_min, rel_humid_max, temp_med, rel_humid_min |
| Rio Grande do Norte (RN) | 24 | rel_humid_med, thermal_range, rel_humid_max, temp_max, rel_humid_min |
| Rondônia (RO) | 43 | pressure_min, rel_humid_min, pressure_max, rel_humid_med, rainy_days |
| Roraima (RR) | 11 | rainy_days, temp_med, rel_humid_max, pressure_max, precip_max |
| Rio Grande do Sul (RS) | 14 | rel_humid_max, temp_med, rel_humid_med, temp_min, pressure_med |
| Santa Catarina (SC) | 35 | rel_humid_max, rainy_days, pressure_min, precip_med, temp_min |
| Sergipe (SE) | 42 | pressure_min, pressure_med, pressure_max, rainy_days, thermal_range |
| São Paulo (SP) | 28 | rel_humid_min, pressure_min, precip_med, temp_max, rel_humid_med |
| Tocantins (TO) | 17 | rel_humid_max, temp_med, precip_min, thermal_range, rel_humid_med |

**Fig. 9.**
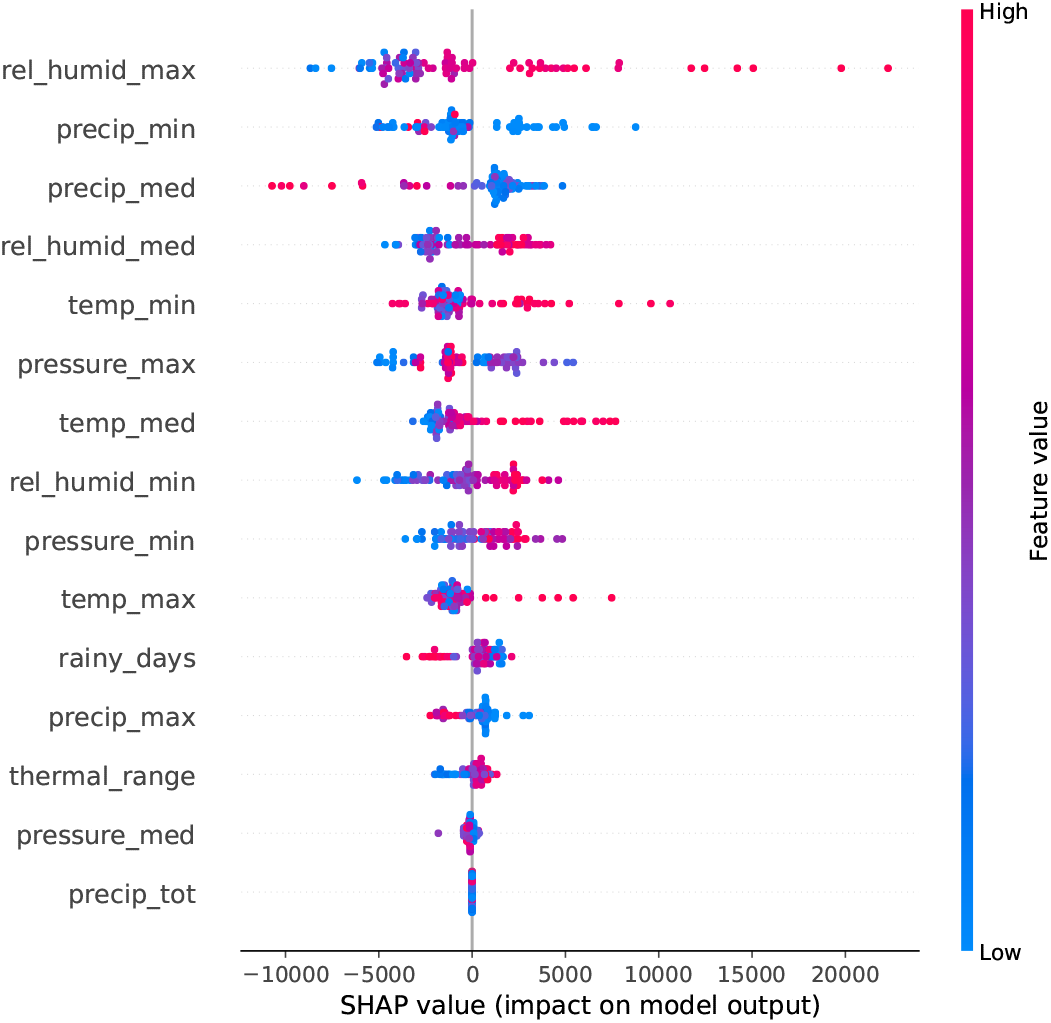
SHAP feature importance for dengue case prediction in Minas Gerais, Brazil.

### 3.2 Model performance

In this section, we present the performance of the four models evaluated:

1. LSTM-Cases: LSTM using only dengue case data.

2. LSTM-Climate: LSTM with lagged climate variables and seasonal patterns.

3. LSTM-Climate-Spatial: LSTM with lagged climate variables, seasonal patterns, and neighboring spatial effects.

4. Bayesian Baseline: Bayesian random effects model using the same covariates as Model 3.

Tables 4 and 5 present the forecasting performance of the models for 1-month and 3-month prediction horizons, respectively. As shown in these tables, in general our proposed LSTM-Climate-Spatial model achieves superior performance across the majority of states, outperforming the other models in terms of MAE, MAPE, and CRPS. This underscores the effectiveness of combining lagged climate variables, seasonal patterns, and spatial effects in capturing the dynamics of dengue transmission. Additionally, Figures S1 and S2 in the supplementary materials show the time series plots of the forecasts for all 27 states across the four models for the 1-month and 3-month horizons, respectively. These figures provide a detailed visualization of the models’ performance over time, highlighting the differences in prediction accuracy across the states.

**Table 4.**
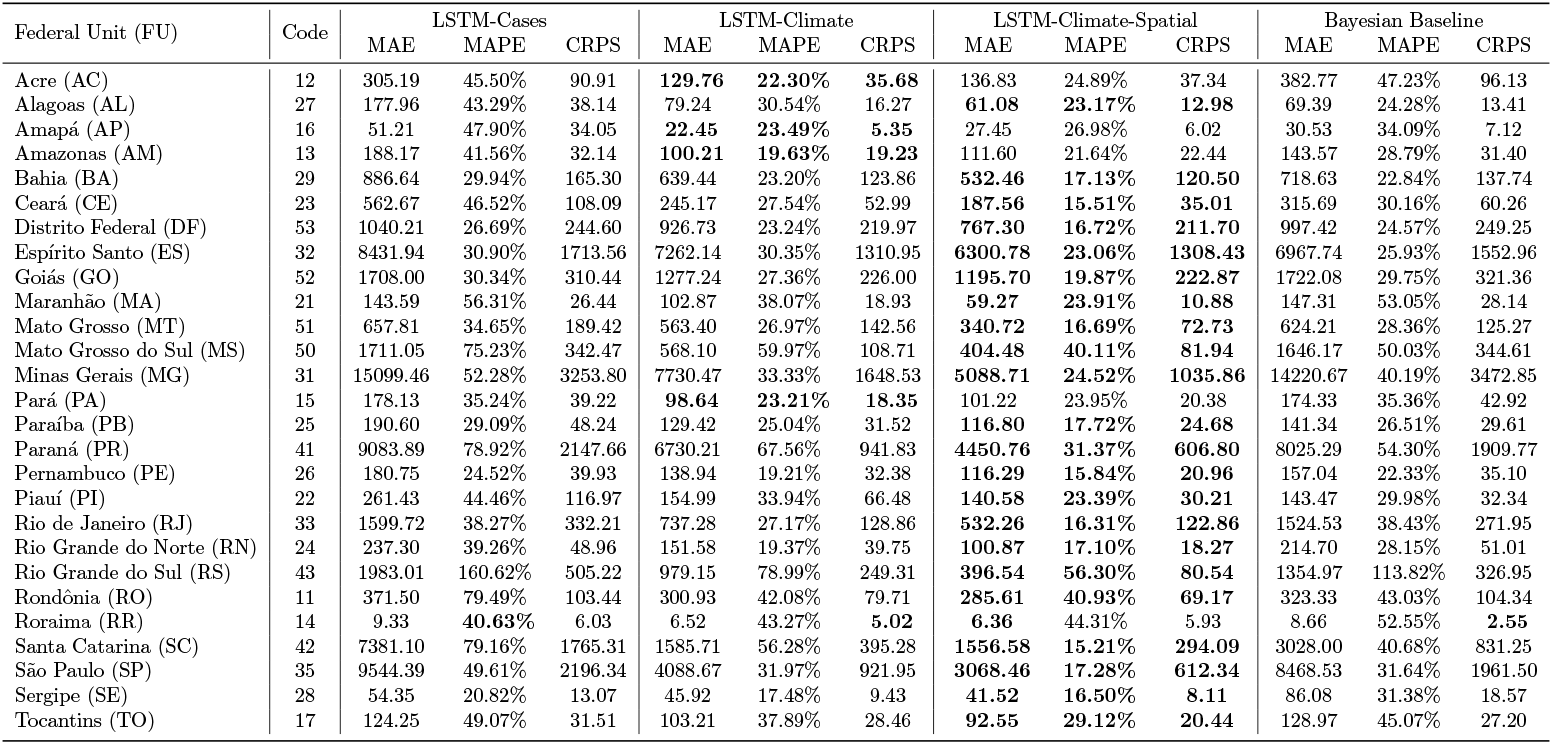
Forecasting performance for 1-month horizon across Brazilian states using different models.

**Table 5.**
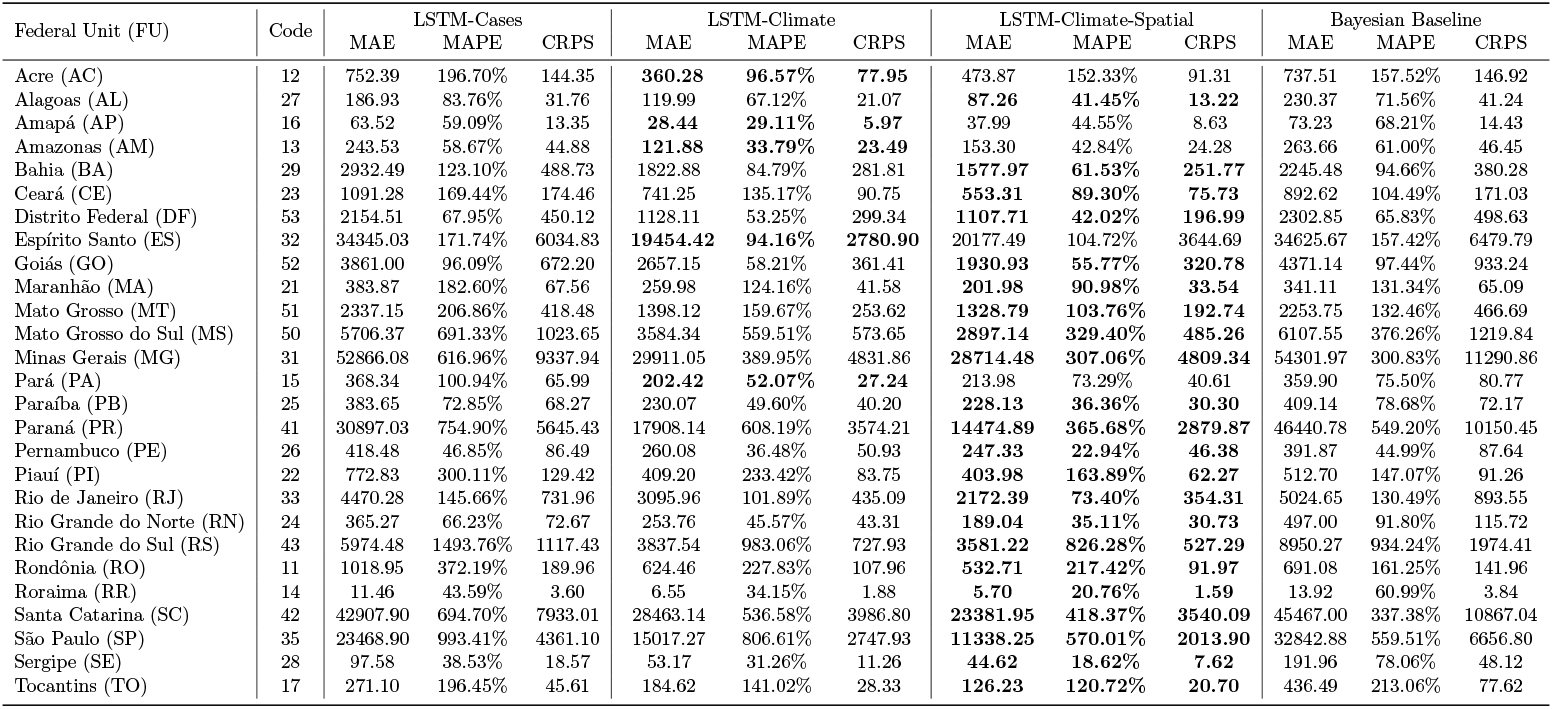
Forecasting performance for 3-month horizon across Brazilian states using different models.

In most states, the inclusion of spatial dependencies through neighboring state dengue data improves model accuracy, particularly in states with high connectivity or where regional influences play a significant role in disease spread. For instance, in Minas Gerais (MG), the integration of spatial effects significantly reduces prediction errors. For 1-month forecasts, the LSTM-Climate-Spatial model achieves an MAE of 5088.71, MAPE of 24.52%, and CRPS of 1035.86, compared to the LSTM-Climate model’s MAE of 7730.47, MAPE of 33.33%, and CRPS of 1648.53. A similar trend is observed for 3-month forecasts, where the LSTM-Climate-Spatial model demonstrates its superiority across all metrics. This makes the LSTM-Climate-Spatial model the most accurate option for forecasting in this geographically diverse state.

Similar improvements are observed in Paraná (PR) and Ceará (CE). In Paraná, the LSTM-Climate-Spatial model achieves a significant reduction in MAE (4450.76 vs. 6730.21 in the LSTM-Climate model for 1-month predictions), as well as improvements in MAPE and CRPS. In Ceará, the inclusion of spatial effects enhances the model’s ability to account for regional transmission dynamics, driven by population movement and climatic factors, with substantial gains in prediction accuracy across all metrics. Goiás (GO) also presents a compelling case where the spatial effects contribute substantially to the model’s success. For 1-month predictions, the LSTM-Climate-Spatial model achieves an MAE of 1195.70, MAPE of 19.87%, and CRPS of 222.87, compared to the LSTM-Climate model’s MAE of 1277.24, MAPE of 27.36%, and CRPS of 226.00. This improvement reflects the considerable connectivity of Goiás with its neighboring states, allowing the model to capture the influence of dengue incidence in surrounding regions. For 3-month forecasts, the trend persists, highlighting the adaptability of the LSTM-Climate-Spatial model to varying spatial structures.

Compared to the Bayesian baseline model, the LSTM-Climate-Spatial model consistently demonstrates better accuracy across most states. For example, for 1-month predictions, in Ceará (CE), the Bayesian baseline model achieves an MAE of 315.69, MAPE of 30.16%, and CRPS of 60.26, while the LSTM-Climate-Spatial model outperforms it with an MAE of 187.56, MAPE of 15.51%, and CRPS of 35.01. Similarly, in Paraná (PR), the Bayesian baseline model shows an MAE of 8025.29 and CRPS of 1909.77, significantly higher than the LSTM-Climate-Spatial’s MAE of 4450.76 and CRPS of 606.80. This highlights the limitations of the Bayesian approach, which struggles to account for the complex temporal and spatial patterns captured effectively by the LSTM-based models.

However, not all states benefit equally from the inclusion of spatial effects. In some cases, the addition of neighboring state data leads to a slight decline in model performance. These states, highlighted in Figure 10, are predominantly located in the northern region of Brazil, including Acre (AC), Amazonas (AM), Amapá (AP), Rondônia (RO), Roraima (RR), and Pará (PA). These states are characterized by vast tropical rainforests and tropical monsoons (Figure 5), which limit population mobility and regional connectivity compared to the more densely populated and urbanized central-west and southeast regions. The relatively sparse population distribution and lower human movement across these areas reduce the influence of spatial dependencies, thereby diminishing the utility of neighboring state dengue case data for improving forecasts.

**Fig. 10.**
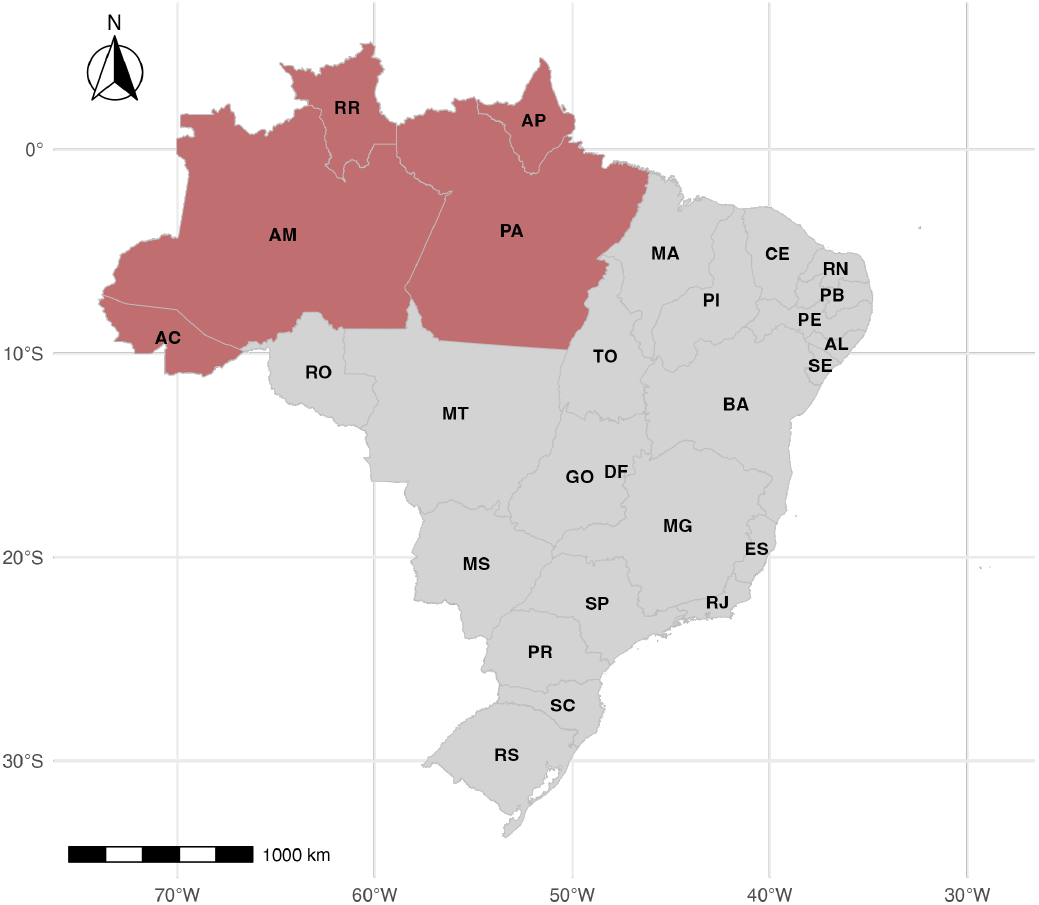
Northern states in Brazil without neighbor influence for dengue forecasting marked in red.

For example, in Acre (AC), the LSTM-Climate-Spatial model shows a slightly higher MAE (136.83) compared to the LSTM-Climate model (129.76) for 1-month predictions. Similarly, in Pará (PA), the MAE increases from 98.64 in the LSTM-Climate model to 101.22 in the LSTM-Climate-Spatial model. This pattern is consistent across CRPS values as well, with AC increasing from 35.68 to 37.34 and PA increasing from 18.35 to 20.38. These results suggest that the inclusion of spatial effects introduces additional noise rather than improving the accuracy of the forecasts in these northern regions.

Despite these limitations, incorporating climate variables still significantly improves forecasting accuracy over models that rely solely on dengue case data. Even in states where spatial effects fail to enhance performance, the LSTM-Climate model consistently outperforms the LSTM-Cases model. For instance, for the 1-month forecast in Amazonas (AM), the MAE for the LSTM-Climate model is 100.21, compared to 188.17 for the LSTM-Cases model, and the CRPS equal to 19.23 improves from 32.14. Similarly, in Pará (PA), the MAE drops dramatically from 178.13 in the LSTM-Cases model to 98.64 in the LSTM-Climate model, and the CRPS improves from 39.22 to 18.35. These findings highlight the robustness of using lagged climate variables to enhance dengue forecasting, even in geographically isolated states where spatial interactions may be less relevant. By relying on climatic conditions that directly affect mosquito breeding and transmission dynamics, the LSTM-Climate model ensures a significant improvement in forecasting accuracy with respect the model that just uses cases data, reinforcing the critical role of environmental factors in dengue prediction.

In summary, the results emphasize the effectiveness of our proposed LSTM-Climate-Spatial model, which achieves the best performance across most Brazilian states. By effectively integrating temporal patterns, lagged climate variables, and spatial dependencies, this model captures the complex dynamics of dengue transmission across diverse regions. The careful selection of climate variables, guided by SHAP, ensures that the model captures critical climatic factors driving dengue transmission. For the majority of states, the integration of spatial effects further enhances forecasting accuracy, making the LSTM-Climate-Spatial model the most robust and reliable approach. Even in a few isolated states, such as Acre and Roraima, where spatial effects provide limited benefits, the climate component still significantly outperforms models relying solely on dengue case data. These results underscore the robustness and adaptability of the LSTM-Climate-Spatial model, reinforcing its critical role in improving dengue forecasting accuracy across diverse epidemiological and geographic contexts in Brazil, and establishing it as a reliable tool for medium-term dengue forecasting and a valuable asset for public health planning across Brazil.

## 4 Discussion

This study addresses the challenges of dengue forecasting at a national scale by developing a robust model capable of integrating climatic, temporal, and spatial information. Unlike many previous studies that focus on specific cities or states or employ models that fail to fully capture spatial-temporal dynamics, our work presents a comprehensive LSTM-based framework applied to all 27 Brazilian states. By incorporating lagged climate variables, seasonal patterns, and spatial effects from neighboring states, our approach provides a scalable and adaptable solution for forecasting dengue cases across diverse geographic and climatic contexts.

The proposed LSTM-Climate-Spatial model consistently outperforms baseline models, demonstrating its ability to capture the multifaceted dynamics driving dengue transmission. This superiority stems from several key innovations. First, the inclusion of SHAP-selected climate variables ensures that only the most relevant climatic factors are utilized, enhancing the model’s predictive power. Second, the integration of spatial dependencies leverages cross-regional influences, effectively capturing transmission patterns in states such as Minas Gerais, Paraná, Ceará, and Goiás. These states, located predominantly in the Southeast and Midwest regions, are characterized by higher population densities and greater human mobility, compared to the northern states. Furthermore, the climates prevalent in these regions facilitate favorable conditions for dengue transmission, increasing the interconnectedness of dengue dynamics across state borders. These features make the model not only highly accurate but also adaptable to the heterogeneous transmission dynamics observed across Brazil.

Additionally, the use of lagged climate data allows the model to account for delayed effects of climate factors on dengue transmission, further improving its predictive reliability. The incorporation of temporal patterns, such as seasonal cycles, enables the model to adapt to recurring trends, ensuring robust performance across short- and medium-term forecasting horizons. These elements collectively make the LSTM-Climate-Spatial model a highly effective tool for understanding and forecasting dengue, providing valuable insights for public health planning and intervention strategies. The selected lags (1–3 months for predicting dengue cases 4 weeks ahead and 3 months for predicting 12 weeks ahead) align with previous studies, and also reflect data availability since climate data is typically available only up to the current time. These lags capture both immediate and cumulative climatic effects on dengue transmission by accounting for the timing of environmental influences on mosquito dynamics. In specific public health settings, the lag duration can be appropriately adjusted based on local conditions and the availability of real-time climate data to ensure the most accurate and actionable forecasts.

While our approach shows significant promise, challenges remain. One notable limitation is the variability in spatial effects across states, which underscores the need for a deeper exploration of region-specific factors. The influence of spatial dependencies can be shaped by diverse elements, such as human mobility patterns, socio-economic factors, vector ecology, and local epidemiology [61–63]. These factors are often highly heterogeneous and can vary not only between states but also within regions, affecting the ability of spatial models to capture dengue transmission dynamics accurately.

Our current model accounts for spatial effects by incorporating dengue cases from neighboring states, where neighboring is defined as sharing a physical border. While this approach provides a useful baseline, it is somewhat simplistic and may not fully represent the complex interconnectivity between regions [64]. For example, human mobility does not always align with geographic borders, and highly connected urban centers or transportation hubs might influence dengue transmission across greater distances or through indirect pathways. Future work could consider more advanced methods to capture spatial effects, such as integrating mobility data, air travel patterns, or broader regional interactions beyond direct neighbors. These enhancements would allow the model to better reflect the intricate spatial dynamics that influence dengue outbreaks.

## 5 Conclusion

This study presents a comprehensive and scalable framework for dengue forecasting across Brazil, integrating temporal, climatic, and spatial information into an LSTM-based model. Our results reveal that the proposed LSTM-Climate-Spatial model consistently outperforms baseline models in most states, underscoring its ability to capture complex dynamics in dengue transmission.

The incorporation of SHAP-selected climate variables proves to be a pivotal improvement, ensuring that the model focuses on the most influential predictors, which directly reflect climatic drivers of dengue outbreaks. The inclusion of spatial effects also significantly enhances model performance in states with strong inter-regional connectivity, such as Minas Gerais, Paraná, Ceará, and Goiás. This demonstrates the utility of leveraging neighboring state data to capture cross-regional transmission patterns. However, in geographically isolated states like Acre and Roraima, where inter-regional interactions are limited, spatial effects introduced noise and slightly reduced model accuracy. Nonetheless, even in these cases, the climate-enhanced models consistently outperformed case-only models, reaffirming the critical role of climate variables in dengue forecasting.

This work represents a significant step toward national-scale dengue forecasting in Brazil, addressing gaps in previous studies that often focus on limited regions or lack the integration of spatial-temporal dynamics. Future research should explore more sophisticated methods to model spatial dependencies, such as incorporating human mobility data or broader regional interactions, to further enhance prediction accuracy. Despite these challenges, the results underscore the utility of combining climate, spatial, and temporal data for robust dengue forecasting, providing valuable insights to inform public health strategies and outbreak preparedness in Brazil.

## Data Availability

All data produced are available online

## Supplementary information

**Fig. S1.**
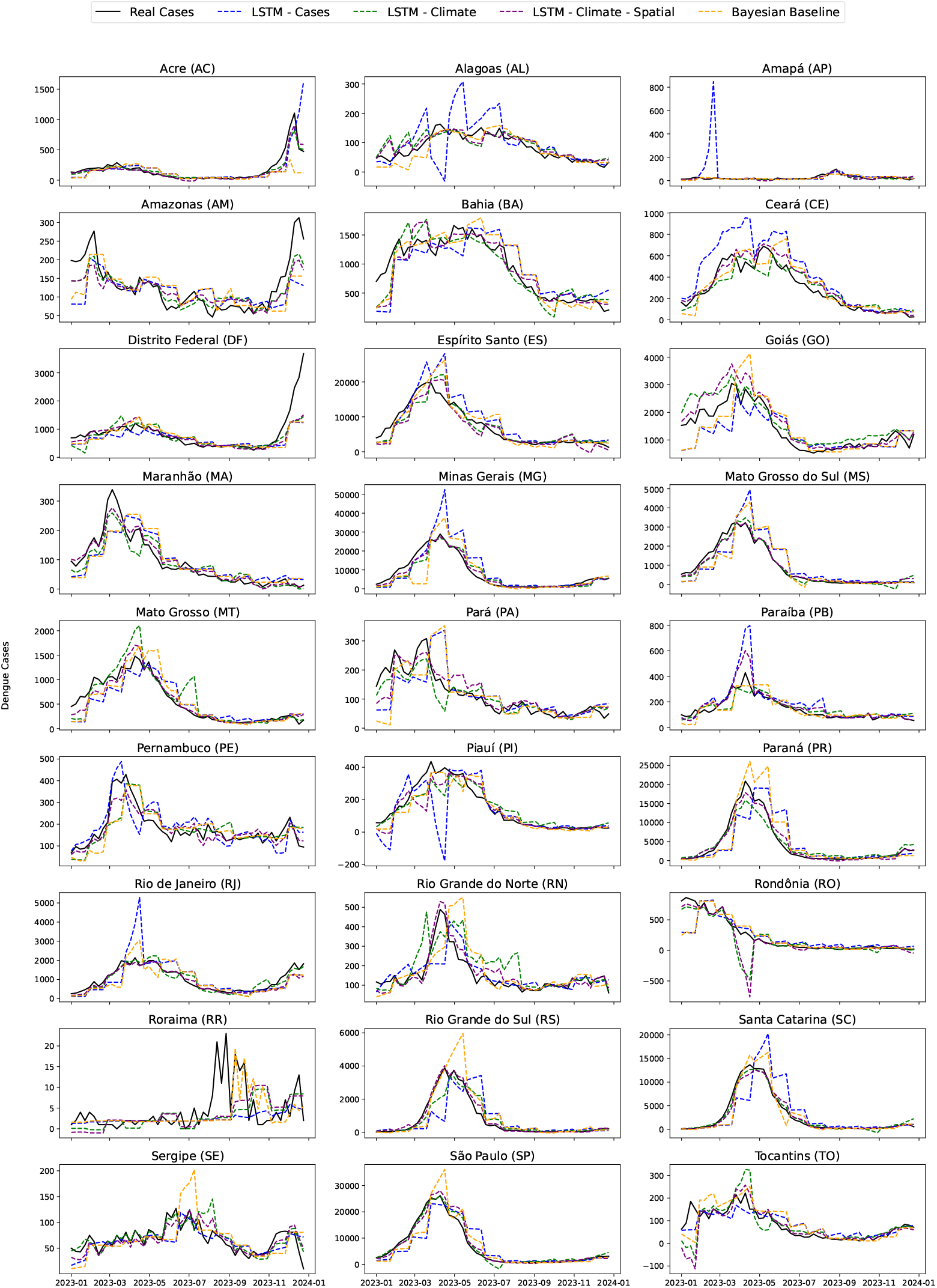
Observed dengue cases and model forecasts for 1-month horizon across 27 Brazilian states.

**Fig. S2.**
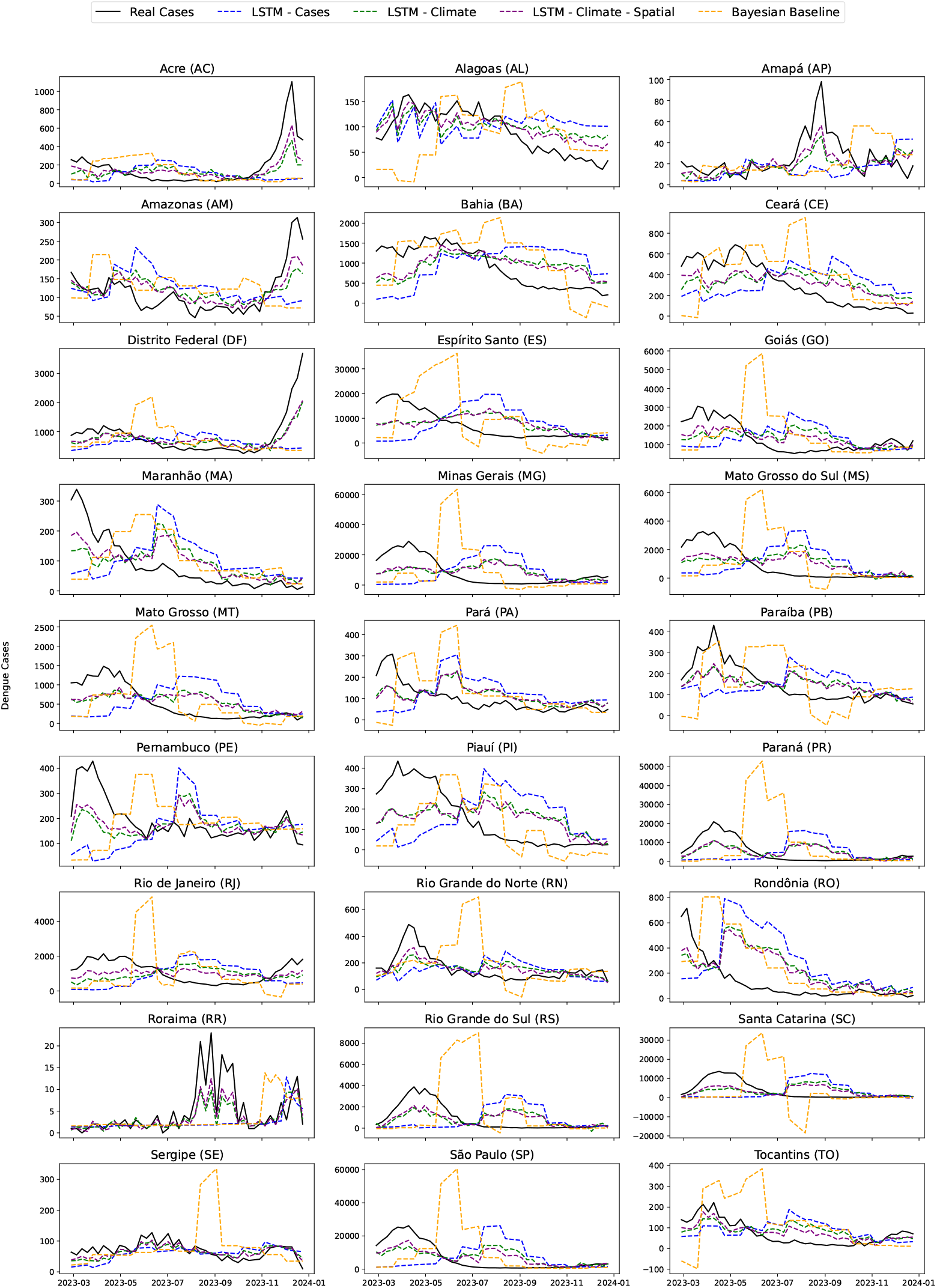
Observed dengue cases and model forecasts for 3-month horizon across 27 Brazilian states.

## Declarations

### Funding

This research received financial support from The Letten Prize (https://lettenprize.com/), with a personal award to Paula Moraga. The funders had no role in study design, data collection and analysis, decision to publish, or preparation of the manuscript.

### Competing interests

The authors declare that they have no competing interests.

### Availability of data and materials

All data used in this study is open access and freely available on the internet, see the “Methods” section for details. For reproducibility purposes, we provide a permanent GitHub repository with the codes, which can be found at https://github.com/ChenXiang1998/LSTM-Based-Dengue-Prediction-Across-Brazil.

### Author contribution

PM and XC conceived and designed the study. XC collected and processed the data, and performed the experiments. All the figures and maps are drawn by XC. XC and PM were contributors in writing and revising the manuscript. All authors read and approved the final manuscript.

### Ethics approval and consent to participate

Not applicable.

### Consent for publication

Agreement for publication.

## Notes

### Competing Interest Statement

The authors have declared no competing interest.

